# Influence of sex hormone use on sleep architecture in a transgender cohort: findings from the prospective RESTED study

**DOI:** 10.1101/2023.06.22.23291701

**Authors:** Margot W. L. Morssinkhof, Ysbrand D. van der Werf, Odile A. van den Heuvel, Daan A. van den Ende, Karin van der Tuuk, Martin den Heijer, Birit F. P. Broekman

## Abstract

Sex differences in sleep architecture are well-documented, with females experiencing longer total sleep time (TST), more slow wave sleep (SWS) and shorter Rapid Eye Movement (REM) sleep duration than males. Although studies imply that sex hormones could affect sleep, effects of exogenous sex hormones on sleep architecture remain unclear. This study examined sleep architecture changes in transgender individuals after 3 months of gender-affirming hormone therapy (GAHT).

We assessed sleep architecture in 73 transgender individuals: 38 transmasculine participants who started using testosterone and 35 transfeminine participants who started using estrogens and anti-androgens. Sleep architecture was measured before GAHT and after 3 months of GAHT for 7 nights using an ambulatory single-electrode sleep EEG device. Changes in sleep architecture were analyzed using linear mixed models, and non-normally distributed outcomes were log-transformed and reported as percentages.

In transmasculine participants, SWS decreased by 7 minutes (95% CI: −12; −3) and 1.7% (95% CI: −3%; - 0.5%), REM sleep latency decreased by 39% (95% CI: −52%; −22%) and REM sleep duration increased by 17 minutes (95% CI: 7; 26) after 3 months of GAHT. In transfeminine participants, sleep architecture showed no significant changes after 3 months of GAHT.

Sleep architecture changes after three months of masculinizing GAHT in line with sleep in cisgender males, while it shows no changes after feminizing GAHT. The sex-specific nature of these changes raises new questions on sex hormones and sleep. Future research should focus on studying possible underlying neural mechanisms and clinical consequences of these changes.

**Statement of significance:** Sleep architecture shows differences between men and women, with women showing longer sleep, longer slow wave sleep and shorter REM sleep than men. Rodent research indicates that sex hormones can alter sleep architecture, but research on sex hormones and sleep architecture in humans is still lacking. This study examined effects of three months of gender-affirming hormone use in transgender individuals. Results show that testosterone use in persons assigned female at birth resulted in sleep architecture changes similar to cisgender males, whereas estradiol- and anti-androgen use by persons assigned male at birth did not change sleep architecture. These novel findings indicate that sex hormones could change sleep architecture in a sex-specific manner, warranting further studies into causal mechanisms underlying these changes.

## Introduction

There are many known physiological differences between men and women, but one of the lesser known sex differences is found in sleep architecture. Sleep architecture, which can be measured with sleep electroencephalography (EEG), refers to the organization of sleep into sleep depth and sleep stages. When measuring sleep with EEG, healthy women generally show better sleep quality than healthy men (e.g. longer total sleep time, less time awake during the night, longer deep sleep and longer REM latency^1, 2^). Although researchers have intensively studied sleep architecture and the underlying neural mechanisms, the cause of these sex differences in sleep architecture is still not fully understood.

One of the hypothesized mechanisms underlying the sex differences in sleep architecture is attributed to the effect of sex hormones. In mice, sex differences in sleep architecture are eliminated after gonadectomy, which results in the loss of sex steroid production^3^. Female mice also show longer REM sleep durations after gonadectomy, and restoration of estradiol levels through supplementation also restores the shorter REM sleep duration ^4, 5^. There are indications that the influence of sex steroids is sex-specific: an experiment using administration of estrogen and testosterone in female and male rats shows that female, but not male, rats show changes in sleep (e.g. increased wakefulness, changes in REM sleep) after administration of either hormone^4^.

In humans, knowledge on sex hormones and sleep architecture is scarce. Studies have mainly focused on clinical groups using hormone therapy, such as women during the perimenopause and hypogonadal men. A few studies in healthy reproductive-age persons showed indications of sex hormone effects on sleep architecture. In healthy men with induced hypogonadism, testosterone supplementation resulted in more deep sleep and a longer REM sleep latency compared to placebo ^6^. Similarly, estrogen therapy in hypogonadal females resulted in decreased sleep onset latency and longer REM sleep duration compared to placebo^7^. Findings on progesterone are less consistent: Progesterone administration was found to have a sedative effect in healthy males and females^8^ but also reduced slow wave sleep duration in males^9^. Administration of a progesterone antagonist in healthy males resulted in increased sleep onset latency and more time spent awake during the night^10^.

One of the major limitations of most studies on effects of sex hormones on sleep is that most studies focus on effects of short-term hormone administration or assessment of effects of sex hormones in persons with hypogonadism, which is known to disrupt sleep^11^. This lack of research makes it difficult to disentangle whether the observed sex differences in sleep architecture in the population are driven by sex hormones, or by other biological sex-specific factors like sex chromosomes or developmental sex differences. One way of gaining insight into the effects of sex hormones on human sleep would be to study the effects of male sex hormones (e.g. testosterone) in females and the effects of female sex hormones (e.g. estrogen) in males, but this is difficult to do in an experimental setting.

Transgender users of gender-affirming hormone therapy (GAHT) use sex hormones to masculinize or feminize their bodies, to better align their physical state with their gender identity ^12^. The study of changes in sleep architecture after GAHT use can offer insight into the direct effect of sex hormone use on sleep architecture. The use of GAHT in transfeminine persons assigned male at birth most often consists of anti-androgens and estrogens which act to feminize the body, and the use of GAHT in transmasculine persons assigned female at birth consists of testosterone use which acts to masculinize the body. Since GAHT use commonly results in sex steroid levels comparable to the opposite sex, this enables us to specifically examine the effects of healthy physiological levels of sex hormones on sleep.

Thus far, the effect of GAHT on sleep architecture is unclear. Only one study examined effects of feminizing GAHT in seven transgender women^13^, and they found that participants show more light sleep after 3 months of estradiol and cyproterone acetate. This study, however, was limited both in sample size and in participant selection, since they only included transgender women.

Therefore, this study aims to examine the changes in sleep architecture after GAHT use. We examine whether characteristics of sleep architecture (e.g. total sleep time, wakefulness after sleep onset, slow wave sleep duration and frequency, number of arousals, number of awakenings, REM sleep duration and REM sleep latency) change after 3 of either feminizing or masculinizing hormone use. We hypothesize that testosterone use in transmasculine persons is associated with shorter total sleep duration, more wakefulness during the night and more arousals and awakenings, shorter slow wave sleep duration and shorter REM sleep latency, and that estrogen and anti-androgen use by transfeminine persons is associated with longer total sleep duration, less wakefulness during the night and less arousals and awakenings, longer slow wave sleep duration and longer REM sleep latency.

## Methods

### Study setting

Data for this study was obtained within the RESTED study (Relationship between Emotions and Sleep in Transgender persons: Endocrinology and Depression), which is an observational prospective study aimed at studying changes in mood and sleep in transgender hormone users during the first year of GAHT. The RESTED study was conducted within the transgender healthcare clinics of the Amsterdam University Medical Center (Amsterdam UMC) and the University Medical Center Groningen (UMCG) and included participants from January 2020 until September 2022. The RESTED study received a declaration stating that Medical Research Involving Human Subjects Act (WMO) did not apply to this data collection (study id: 2019.353). The study was performed in accordance with good clinical practice guidelines and the world medical associations code of ethics (Declaration of Helsinki). All participants provided informed consent for study participation and for use of their medical information for research purposes.

### Participants and measurements

All participants were approached and included before the start of GAHT. Participants were eligible to participate in the RESTED study if they were aged between 18 and 50, if they were diagnosed with gender dysphoria and if they had never used gender-affirming hormones before. They were not included if they had a diagnosed sleep disorder (e.g. obstructive sleep apnea, clinical insomnia or parasomnias), or if they used benzodiazepines or barbiturates at time of the study.

### Gender affirming hormone therapy

All participants started using GAHT after the baseline measurement of the study, and participants were treated with GAHT according to the WPATH guidelines^14^. Transmasculine participants, who were assigned female at birth (AFAB), all started using masculinizing hormones in the form of testosterone. These hormones were either administered via testosterone gel (daily dose of 40.5 mg), testosterone esters (250 mg every 3 weeks) or testosterone undecanoate (1000 mg every 12 weeks). A number of transmasculine participants also used progestins (lynestrenol, 5mg; levonorgestrel IUD, 52mg; medroxyprogesterone, 150mg) or combined hormonal contraceptives, consisting of progestins and estrogens (ethinylestradiol/levonorgestrel, 0.03/0.15 mg; ethinylestradiol/drospirenon 0.02/3 mg) to suppress their menstruation.

Transfeminine participants, who were assigned male at birth, started using feminizing hormones, consisting of anti-androgens, either in the form of oral cyproterone acetate (CPA; daily dose of 10 mg) or intramuscular injections of gonadotropin-releasing hormone analogues (GnRH analogues), either short-working (triptorelin 3.75 mg per 4 weeks, or leuproreline, 3.75 mg per 4 weeks) or long-working (triptorelin 11.25mg per 12 weeks), combined with estradiol, which was either administered orally (estradiol valerate, 2mg twice daily) or transdermally through estradiol gel (0.06% 1.5 mg daily) or estradiol patches (100 mcg per 24 hours, twice a week). For the current study, transfeminine participants were excluded if they only started using estrogens (n = 1) or anti-androgens (n = 1) but not both at the start of GAHT.

### Demographic and clinical characteristics

To collect information on participants’ demographic characteristics, data from electronic patient files was combined with data from study surveys. At each outpatient clinic appointment participants were asked about medication use and intoxications (e.g. smoking, alcohol and drug use). As part of regular care, presence of possible psychiatric diagnoses was tested using the MINI+ (Mini International Neuropsychiatric Interview^15^), a structured clinical interview, at the intake appointment of the clinic. In both participating centers, serum hormone levels were measured as part of regular care at the start of GAHT use, after 3 months of GAHT use and after 12 months of GAHT use. In both the Amsterdam UMC and the UMCG, serum testosterone measurements were conducted using liquid chromatography tandem mass spectrometry (LC-MS/MS) with a lower limit of quantitation of 0.1 nmol/L, and an inter-assay coefficient of variation of 4% to 9%. Serum estradiol measurements in both centers were conducted using LC-MS/MS with a lower limit of quantitation of 20 pmol/L and an inter-assay coefficient of variation of <7%.

### Sleep measurements

The study consisted of three periods of sleep measurements of one week each: The first measurement was conducted before the start of GAHT, the second measurement after 3 months of GAHT, and the third measurement after 12 months of GAHT. In each measurement week, participants were asked to record sleep architecture during seven nights using an ambulatory single-electrode EEG sleep measurement device (Smartsleep, Philips, the Netherlands). Subjective sleep was measured with the consensus sleep diary^16^ and self-reporting questionnaires on sleep (the Pittsburgh Sleep Quality Index^17^, or PSQI, and Insomnia Severity Index^18^, or ISI).

#### Sleep diaries and sleep questionnaires

The consensus sleep diary^16^ (CSD) is a standardized diary to track information on daily sleep patterns, including sleep-specific questions about time spent in bed, total sleep time, sleep onset latency and amount of times and duration of wakefulness during the night. The CSD also asks about factors affecting sleep, such as coffee consumption, alcohol consumption and use of substances or medication that affect sleep. For the current study, questions were added to the CSD which inquired about experiences using the sleep tracker. A comment field was added in which participants could note comments about the measurement night, including remarks on the sleep tracker.

To assess sleep quality and symptoms of insomnia, the Pittsburgh Sleep Quality Index (PSQI) and Insomnia Severity Index (ISI) were used at every measurement. The PSQI is a 19-item questionnaire that broadly assesses components of sleep quality, including sleep duration, sleep quality, sleep disturbances and daily burden of sleep problems^17^. It is scored into seven subscores, which are then added up to form a sum score (range 0 to 21). The PSQI has an established cutoff score of 5, with a score of 5 or lower indicating “good” sleep and a score higher than 5 indicating “poor” sleep. The ISI is a seven-item questionnaire that inquires about symptoms of insomnia in the previous two weeks (Morin *et al.,* 1993). The seven items are all scored from 0 to 4, resulting in a sum score from 0 to 28. There are clinical cutoffs for the ISI scores, with a score below 7 indicating no presence of insomnia, 8 to 14 indicating subclinical insomnia, 15 to 21 indicating clinical insomnia and a score over 21 indicating severe insomnia. The items in the ISI correspond to the DSM-5 defined criteria for clinical insomnia, and is a widely used tool to assess the burden of insomnia symptoms. To assess perceived stress and depressive symptoms, the Perceived Stress Scale^19^ (PSS) and Inventory of Depressive Symptomatology-Self Report^20^ (IDS-SR) were used. Questionnaire results for both were used to assess the presence of selection bias in the sleep EEG measurements as displayed in the supplementary materials in Table S2.

#### Sleep architecture

Sleep architecture, consisting of Sleep Onset Latency (SOL), Total Sleep Time (TST), Wake After Sleep Onset (WASO), Sleep Efficiency (SE), Slow Wave Sleep duration (SWS), Number of Arousals (NRA; < 5 minutes awake), Number of Interruptions (NRI; > 5 minutes awake), REM sleep duration and REM sleep latency was measured using the EEG sleep measurement device. This sleep measurement device uses dry single-lead EEG for measuring sleep, with electrodes at Fpz and M1. This EEG signal can differentiate wakefulness, light sleep (i.e. phase 1 and 2), deep sleep (i.e. phase 3 sleep or SWS) and REM sleep. Participants were asked to record their sleep at home. After completing a week of measurements, the device was brought or sent back to the hospital and the data on the device was uploaded to the study database by the local research team.

Measurements were cleaned to exclude incomplete nights or nights with poor measurement quality: a measurement night was considered incomplete if the participant reported that they took off the headband during the night in the sleep diary, if the measured TST, WASO and SOL together were shorter than 240 minutes or if the contact impedance was higher than the devices’ internal threshold limit threshold for good quality recordings. For the analyses, the first night of each sleep architecture measurement week was not used for analysis, since the first night was considered a habituation night. The SE was calculated by dividing the TST by the total duration of the sleep episode (e.g. the sum of the TST, SOL and WASO) and multiplying this ratio by 100 to get a percentage.

In 26 of the 121 measurements, the device was inadvertently set in active mode (as described in Garcia-Molina *et al*., 2018^21^). The active mode is designed to increase slow wave amplitude in sleep restricted populations, but was shown to not affect sleep architecture (e.g. total sleep duration, WASO, SWS) (Garcia-Molina *et al.,* 2018). Sensitivity analyses incorporating device setting as covariate show that the active mode did not significantly affect measurements, as shown in the supplementary materials 2. Hence, upon consideration, we deemed this not to be a significant factor for our research questions.

### Statistical analyses

Continuous outcomes were summarized with means and standard deviations (SDs) if they were normally distributed and medians, 25^th^ percentile and 75^th^ percentile if they were not normally distributed. Categorical outcomes were summarized based on counts and percentages. Outcome variables with a non-normal distribution in residuals were log-transformed to ensure that the data met the assumptions of the statistical analyses.

Changes in sleep architecture were analyzed using linear mixed models in Rstudio (version 1.3.1093) using packages “lme4”^22^ and lmerTest^23^. To estimate changes in sleep architecture after 3 months of GAHT, a model was constructed with the sleep outcomes per night (SOL, TST, WASO, SE, NRI, NRA, SWS, SWS%, REM sleep latency and REM sleep duration) as outcome variables, timepoint (e.g. baseline or 3 months of GAHT) as the fixed predictor and a random intercept per participant to adjust for repeated measures within participants. Since a number of participants reported the use of psychotropic medication, we conducted an additional sensitivity analysis in which we adjusted for the use of psychotropic medication by adding this as a covariate in the model. To correct for repeated testing of the main results, Bonferroni correction was applied to all the main analyses, resulting in a corrected p-value threshold of 0.0025.

## Results

### Demographic and clinical characteristics

Figure 1 displays the sample size of the full RESTED study and the sample size of the current study. The total study sample of the current study consists of 38 transmasculine participants, of whom 36 contributed data in the baseline measurement and 26 contributed data at the 3-month follow-up, and 35 transfeminine participants, of whom 32 contributed data to the baseline measurement and 24 contributed data to the 3-month follow-up measurement. In order to use the available data in the most optimal manner, all available data was used, including data from participants whose data was only present in one of the two measurements. As displayed in table 1, transmasculine participants’ median age at baseline was 23 and transfeminine participants’ median age at baseline was 26. At baseline, 17% of transmasculine and 12% of transfeminine participants used psychotropic medication, most commonly antidepressants (11% in transmasculine and 9% in transfeminine participants) and stimulants (6% in transmasculine and 6% in transfeminine participants).

**Figure 1.**
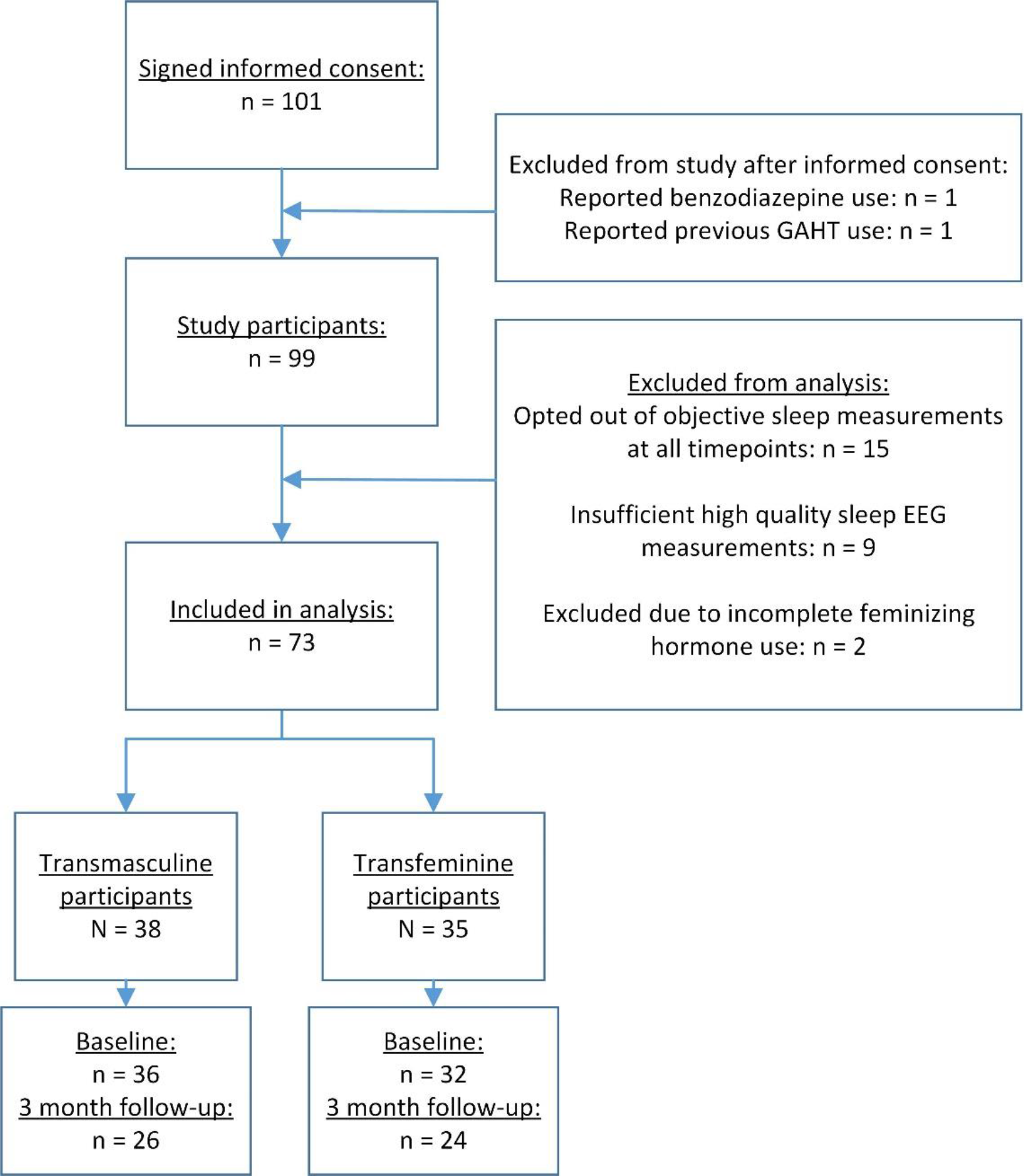
Inclusion flowchart of the RESTED study, showing inclusion rates and sample sizes per group and timepoint in the study.

**Table 1.** Demographic and clinical characteristics at the baseline measurement and 3 month follow up.

|  |  |  |  |  |  |
| --- | --- | --- | --- | --- | --- |
|  |  | Transmasculine participants (n = 38 ) |  | Transfeminine participants (n = 35) |  |
| Measurement |  | Baseline<br>n = 36 | 3 months<br>n = 26 | Baseline<br>n = 32 | 3 months<br>n = 24 |
| Age | median, 25<sup>th</sup> and 75<sup>th</sup> percentile | 23 (21 to 25) | 22 (20 to 24.8) | 26 (23.8 to 29.3) | 27 (24 to 33) |
| Center (n, %) | Amsterdam UMC | 31 (86.1%) | 23 (88.4%) | 29 (90.6%) | 20 (83.3%) |
|  | UMCG | 5 (13.9%) | 3 (11.5%) | 3 (9.4%) | 4 (16.7%) |
| Psychotropic medication use (n, %) | Any psychotropic medication | 6 (16.7%) | 5 (19.2%) | 4 (12.5%) | 4 (16.7%) |
|  | Antidepressants | 4 (11.1%) | 3 (11.5%) | 3 (9.4%) | 1 (4.2%) |
|  | Antipsychotics | 1 (2.8%) | 0 (0%) | 0 (0%) | 0 (0%) |
|  | Anxiolytics | 0 (0%) | 0 (0%) | 0 (0%) | 0 (0%) |
|  | Stimulants | 2 (5.6%) | 2 (8.3%) | 2 (6.3%) | 2 (8.3%) |
|  | Mood stabilizers | 0 (0%) | 0 (0%) | 0 (0%) | 0 (0%) |
| Testosterone serum level (nmol/L; median, IQR) <sup>1.</sup> | No cycle regulation use (in TM participants) | 0.9 (0.8 to 1.2) | 16 (9 to 23) | 14 (10 to 17) | 0.6 (0.4 to 0.8) |
|  | Cycle regulation use (in TM participants) | 0.6 (0.4 to 0.9) | 15 (12 to 18) | - | - |
| Estradiol serum level (pmol/L, median; IQR) <sup>1.</sup> | No cycle regulation use (in TM participants) | 193 (135 to 291) | 153 (109 to 202) | 76 (63 to 90) | 280 (177 to 378) |
|  | Cycle regulation use (in TM participants) | 77 (46 to 104) | 99 (71 to 166) | - | - |
| PSQI score | median, 25<sup>th</sup> and 75<sup>th</sup> percentile | 7 (3.5 to 9.5) | 5 (4 to 7) | 6 (4 to 7) | 5 (3.5 to 7) |
| ISI score | median, 25<sup>th</sup> and 75<sup>th</sup> percentile | 6 (4.5 to 10) | 6 (4 to 7.5) | 5.5 (3 to 9) | 5.5 (3 to 10.3) |
| <sup>1.</sup> Reference ranges in cisgender persons: testosterone 9 nmol/L to 30 nmol/L and estradiol 12 pmol/L to 126 pmol/L in cisgender males and testosterone 0.3 to 1.6 nmol/L and 31 pmol/L to 2864 pmol/L in cisgender females (Amsterdam UMC Endocrinologisch Lab, z.d.; Verdonk et al., 2019).<br>According to WPATH SOC 8 guidelines, treatment providers should aim for serum levels associated with users' gender identity, e.g. transmasculine GAHT users meeting the cis male reference range and transfeminine GAHT users meeting the cis female reference range. |  |  |  |  |  |

At baseline, 50% of transmasculine participants used a form of cycle regulation: 11.1% used progestin-only oral forms, 22.2% used progestin-only non-oral forms, and 16.7% used a combined oral contraceptive with estradiol and progestins. After starting GAHT use, 89% of the transmasculine participants used testosterone in the form of testosterone gel, and 11% used testosterone in the form of short-acting testosterone ester injections. In the transfeminine participants after GAHT start, 53% used estradiol in the form of oral estradiol tablets, 38% used transdermal estradiol patches and 9% used transdermal estradiol gel. All transfeminine participants used combined estradiol use with anti-androgen use, with 84% of transfeminine participants using short-acting triptorelin injections, 9.4% using short-acting leuproreline injections and 9.4% using oral cyproterone acetate. All GAHT forms at each follow-up are also reported in Table S1 (see supplementary materials).

### Changes in sleep architecture after GAHT

#### Transmasculine participants

A total of 38 transmasculine participants were included in the data of the current study: 36 participants contributed 146 measurement nights to the baseline measurement, and 26 participants contributed 118 measurement nights to the 3-month follow-up measurement. As shown in table 2, statistical analyses show that after 3 months of GAHT, results show a trend towards a decreased sleep onset latency (−20.5%, p = 0.051), no significant change in the total sleep time and a trend towards a decrease in wake after sleep onset (−14.2%, p = 0.085). After 3 months of GAHT, the number of interruptions (NRI) showed no significant change, but the number of arousals (NRA) decreased (−10.8%, p = 0.044) and the SE improved (2.3%, p = 0.036). Furthermore, after 3 months of GAHT slow wave sleep duration (−7.4 minutes, p = 0.003) and slow wave sleep percentage (−1.7%, p = 0.007) decreased, REM sleep latency decreased (−38.6%, p = 0.0001) and REM sleep duration increased (16.5 minutes, p = 0.0008). Correction for multiple testing showed that the changes in REM sleep latency and REM sleep duration remained significant after correction for repeated testing (p = 0.0025). See figure 2 for visualization of the sleep architecture outcomes.

**Figure 2.**
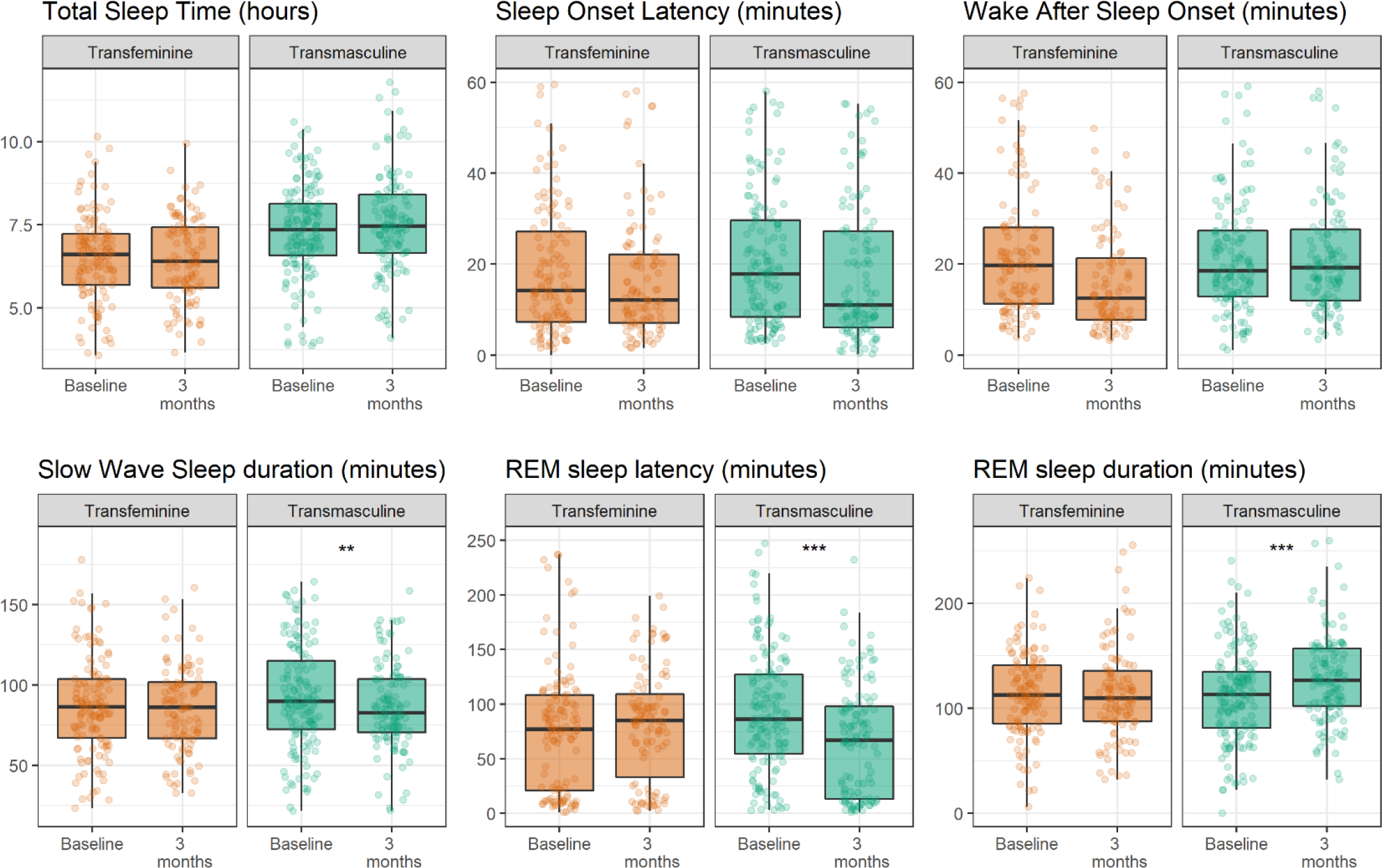
Box plots displaying medians and interquartile ranges (IQRs) of the outcome variables. Dots display the spread of the individual measurement nights; a number of outliers outside of the IQR are not displayed. ** = p < 0.005, *** = p < 0.0005.

**Table 2:** Sleep architecture before GAHT use and after 3 months of GAHT use.

|  | Transmasculine participants |  |  |  | Transfeminine participants |  |  |  |
| --- | --- | --- | --- | --- | --- | --- | --- | --- |
| Timepoint | Baseline | 3 months | Statistical difference<sup>1</sup> |  | Baseline | 3 months | Statistical difference<sup>1</sup> |  |
| n (nights) | 36 (146) | 26 (118) | Unadjusted model | Adjusted model <sup>2</sup> | 32 (124) | 24 (102) | Unadjusted model | Adjusted model <sup>2</sup> |
| **SOL (minutes)** | 19.5 (9.7 to 34.9) | 13.4 (7.0 to 34.6) | -20.5% (37.9% to 0.0%)<br>p = 0.051 | -24.2% (-10.3% to -3.9%)<br>p = 0.023 | 14.97 (7.5 to 30.7) | 13.1 (7.4 to 24.4) | -1.12% (-5.93% to 3.86%)<br>P = 0.65 | -1.11% (-5.9% to 3.8%)<br>p = 0.66 |
| **TST (hours)** | 7.3 (1.41) | 7.53 (1.52) | 0.17 (-0.15 to 0.49)<br>p = 0.29 | 0.20 (-0.12 to 0.53)<br>p = 0.23 | 6.52 (1.32) | 6.47 (1.28) | 0.006 (-0.31 to 0.32) p = 0.97 | 0.005 (-0.30 to 0.32)<br>p = 0.97 |
| **WASO (minutes)** | 21.9 (14.0 to 38.7) | 19.6 (12.6 to 29.7) | -14.2% (-27.9% to 2.1%)<br>P = 0.085 | -14.6% (-28.7% to 2.4%)<br>p = 0.087 | 21.06 (11.7 to 39.7) | 14.98 (8.29 to 31.60) | -8.64% (-24.0% to 9.55%)<br>P = 0.33 | -8.6% (-24.0% to 9.52%)<br>P = 0.34 |
| **SE (%)** | 89.9 (85.6 to 94.5) | 91.9 (85.6 to 94.5) | 2.3% (0.1% to 4.6%)<br>p = 0.044 | 2.5% (0.2% to 4.9%)<br>p = 0.036 | 90.4 (83.6 to 93.6) | 92.4 (86.4 to 95.2) | 0.9% (-2.3% to 2.5%)<br>P = 0.94 | 0.1% (-2.3% to 2.5%)<br>P = 0.95 |
| **NRI (n)** | 1 (0 to 2) | 1 (0 to 2) | -7.7% (-18.8% to 4.9%)<br>P = 0.22 | -8.0% (-19.4% to 5.0%)<br>P = 0.22 | 1 (0 to 2) | 0.5 (0 to 1) | -3.23% (-16.1% to 11.5%)<br>p = 0.65 | -3.13% (-16.1% to 11.5%)<br>p = 0.66 |
| **NRA (n)** | 29 (21 to 44) | 25 (19 to 35.5) | -10.8% (-20.2% to -0.4%)<br>P = 0.044 | -10.4% (-20.0% to 0.4%)<br>P = 0.059 | 28.5 (18 to 41.25) | 24 (17 to 40) | -0.96% (-12.6% to 12.3%)<br>p = 0.88 | -0.96% (-12.7% to 12.3%)<br>p = 0.88 |
| **SWS (minutes)** | 90.6 (34.2) | 85.2 (27) | -7.4 (-12.2 to -2.6)<br>p = 0.0028 | -8.2 (-13.1 to -3.2)<br>p = 0.001 <sup>3</sup> | 87 (30.6) | 84.6 (28.2) | -3.41 (-8.47 to 1.66)<br>P = 0.19 | -3.40 (-8.5 to 1.7)<br>P = 0.19 |
| **% SWS (relative to TST)** | 21.0 (7.8) | 19.5 (7.0) | -1.7 (-3.0 to -0.5)<br>P = 0.0073 | -2.1 (-3.4 to -0.8)<br>p = 0.0017 <sup>3</sup> | 22.75 (8.36) | 22.2 (7.58) | -1.00 (-2.70 to 0.7)<br>P = 0.25 | -0.99 (-2.7 to 0.7)<br>p = 0.25 |
| **REM sleep latency (minutes)** | 87 (58 to 129) | 68 (12.0 to 99.5) | -38.6% (-52.1% to -21.6%)<br>P = 0.0001 <sup>3</sup> | -41.1% (-54.4% to -24.1%)<br>P = 0.00006 <sup>3</sup> | 78 (21.0 to 109.5) | 85 (33 to 109) | 21.51% (-8.60% to 60.8%)<br>P = 0.175 | 21.6% (-8.70% to 60.6%)<br>P = 0.174 |
| **REM sleep duration (minutes)** | 111.3 (44.2) | 128.5 (41.6) | 16.5 (6.9 to 26.0)<br>P = 0.00085 <sup>3</sup> | 18.2 (8.3 to 28.0)<br>P = 0.0004 <sup>3</sup> | 113.92 (42.1) | 113.88 (44.4) | -2.16 (-11.92 to 7.63)<br>P = 0.66 | -2.21 (-11.9 to 7.57)<br>p = 0.66 |

Outcomes at baseline and 3 months are reported in mean and SD if normally distributed and median, 25<sup>th</sup> and 75<sup>th</sup> percentile if not normally distributed. SOL = sleep onset latency, TST = total sleep time, WASO = wake after sleep onset, SE = sleep efficiency, NRI = number of interruptions, NRA = number of arousals, SWS = slow wave sleep, REM = rapid eye movement.
<sup>1</sup> Estimate, 95% confidence interval and p-value are reported. Estimates for variables that were non-normally distributed (e.g. SOL, WASO, SE, NRI, NRA) were log-transformed and are reported in % of change.
<sup>2</sup> Adjusted for the use of psychotropic medication.
<sup>3</sup> Significant after Bonferroni correction for repeated testing (p = 0.0025).

#### Transfeminine participants

A total of 35 transfeminine participants were included in the data of the current study: 32 participants contributed 124 measurement nights to the baseline measurement, and 24 participants contributed 102 nights of measurements to the 3-month follow-up measurement. As shown in table 2, statistical analyses show no significant changes after 3 months of GAHT compared to baseline (see also figure 2).

#### Psychotropic medication

The incorporation of psychotropic medication use as an additional covariate did not significantly affect the direction or magnitude of the results, as displayed in table 2 in the adjusted model results.

## Discussion

This study was the first to prospectively study effects of masculinizing and feminizing sex hormones on sleep architecture in transgender persons. We found that 3 months of masculine sex hormone use resulted in decreased slow wave sleep duration, decreased REM sleep latency and increased REM sleep duration. However, 3 months of feminine sex hormone use showed no significant effects on sleep architecture. The results support our hypothesis that masculinizing hormones in persons assigned female at birth change the sleep architecture towards the opposite sex. However, feminizing hormones in persons assigned male at birth do not change sleep architecture, which does not support our hypothesis.

In the transmasculine participants, the changes in REM sleep latency and duration and in SWS duration are notable. REM sleep latency shifts from a median of 87 to 68 minutes and the REM sleep duration shows an increase from a median of 111 minutes to 128 minutes. Considering a REM sleep duration of 21% to 30% in proportion of total sleep is considered healthy^24^, these strong changes could be clinically relevant. The changes in SWS duration are small and not consistently significant after correction for repeated testing, with a 7-minute decrease in the raw SWS duration (corresponding to a Cohen’s d of 0.16) or 1.7% decrease in SWS percentage (corresponding to a Cohen’s d of 0.23), but very consistent across participants. This shows that the use of testosterone in persons assigned female at birth could affect SWS duration. Considering that the typically healthy range of slow wave sleep is from 16% to 20% ^24^, and participants showed a decrease from 21% to 19.5%, we would deem this change to be of interest for sleep science but of limited clinical meaning.

Our results in the transfeminine participants mostly are in line with previous work conducted by Künzel et al. (2011)^13^. This study assessed the effects of 3 months of feminizing GAHT, specifically estrogen and cyproterone acetate, in seven transgender women. They found no significant changes in most sleep stages, but reported a specific increase in light (N1) sleep, from 33 minutes to 51 minutes. Our current study did not assess light sleep duration, and we could therefore not replicate the finding in N1 sleep. However, this study by Künzel and colleagues found no other changes in sleep architecture, which is in line with our current findings.

It is important to note that, although transmasculine participants were using testosterone, it is not clear whether the androgenic effects of testosterone are also causing the changes in sleep. Testosterone can affect androgenic pathways, as testosterone or via conversion into dihydrotestosterone (DHT), or it be aromatized locally into estradiol and affect estrogen receptors^25^. Previous work showed that in female rats, treatment with testosterone affected sleep, but treatment with DHT, which cannot be aromatized into estradiol, did not ^4^. Therefore, it is also possible that in our participants, the reported effects are not exclusively caused by the androgenic effects of testosterone, but that the administered testosterone was instead converted into estradiol, and that changes in estrogen activity affected sleep architecture.

There is robust evidence that sleep-regulating areas of the brain are sensitive to sex hormones ^26^. Estradiol has been shown to affect the ventrolateral preoptic nucleus (VLPO): this area showed decreased firing rates after estradiol therapy in mice, although changes in sex hormones in male rats had no effect ^27, 28^ and administration of estradiol into the VLPO resulted in increased physical activity in mice ^29^. Furthermore, estradiol could affect the expression of orexin receptors, a sleep-stabilizing steroid. Orexin receptor expression changes with the estrous cycle in female rats as well as after removal and add-back of estradiol in female rats ^30^, as well as after gonadectomy or add-back of androgens in male rats ^31^. Estradiol could also affect the serotoninergic activity in the dorsal raphe nucleus: exposure to estradiol and progestins has been found to alter expression of serotonin receptors^32^. Changes in firing rates in the VLPO and serotonin system could also affect the duration of REM sleep ^33, 34^ and non-REM sleep^35^. Furthermore, sex hormones are known to affect the suprachiasmatic nucleus (SCN)^36^ and the dynamics of diurnal cortisol via the hypothalamus-pituitary-adrenal (HPA) axis^37^. For a full review of neural mechanisms between sex hormones and sleep, see Dorsey *et al.* (2021).

One explanation for the sex discrepancy in our results, showing that transmasculine participants show altered sleep but transfeminine participants do not, could also be found in possible sex differences in sex hormone sensitivity in the brain. The organizational-activational model of sexual differentiation (as posed by Phoenix *et al.,* 1959 ^38^) states that sex hormones can permanently affect neural development (so-called organizational effect), and they can also have a direct effect on neural activity (so-called activational effect). Studies in rodents show that this model might also apply for sex hormone effects on sleep: manipulation of sex hormone exposure during development in rats also alters the sensitivity to sex hormone effects on sleep. Male rats that underwent neonatal castration, resulting in lack of exposure to androgens, were found to show larger sleep changes after treatment with estradiol and progesterone than male rats that underwent endogenous puberty and were therefore exposed to androgens^39^. Vice versa, female rats that were treated with masculinizing postnatal testosterone treatment showed no sleep changes after sex hormone manipulations, whereas the untreated females which were exposed to endogenous puberty and female sex hormone exposure showed significant sleep changes after sex hormone changes ^4^. These findings indicate that exposure to female sex hormones or lack of exposure to male sex hormones could possibly alter the brain’s sensitivity to sex hormone changes.

The current study has a number of strengths. First of all, it was the first prospective study to investigate the effects of both feminizing and masculinizing GAHT on sleep architecture. In our setup, we used a single-electrode EEG device that could be used for ambulatory measurements, which means that participants were able to measure their sleep for multiple days at home. This strengthens the external validity of our sleep measurements, since participants slept in their normal home situations. Furthermore, our study design allows us to examine the effects of longer term hormone use compared to previous work. Many studies assessing effects of sex hormones on sleep architecture focus on the effects of acute-or very short-term hormone exposure^8, 9^, which means results typically display the acute effects, but not the longer term effects of sex hormone use. Compared to these studies, 3 months of GAHT exposure shows the effects of longer-term hormone use. However, many transgender GAHT users will typically use GAHT for the rest of their lives^12^, and future work should focus on longer-term effects of GAHT.

There are also a number of limitations of this study. Firstly, it is possible that there is a form of selection bias in the participant group. Some participants reported that they found the sleep EEG device uncomfortable or that they forgot to charge or to wear the device, and opted out of the measurements. In the supplementary materials, we have supplied an overview of the participant demographics (e.g. age, psychotropic medication use and symptoms of depression, stress or insomnia) of participants who participated in the objective sleep measures compared to those who did not (see Table S3; supplementary materials). These data show no clinically significant differences in symptoms of insomnia, depression or perceived stress between the groups, indicating that the participants who opted in or out of the sleep EEG measurements were comparable. However, it is still possible that the participants with more poor sleep were less likely to participate in the objective sleep measures or less likely to participate in the follow-up measurements altogether.

Secondly, we opted not to conduct full-head sleep polysomnography (PSG) measurements, which means we could not study REM sleep according to the “golden standard” measure, which includes an eye movement electrode, and we could not measure sleep EEG microarchitecture, such as micro-arousals, sleep spindles and eye movement. For further study of sex hormones on sleep, we would also recommend further assessing sleep architecture changes using a multi-channel PSG measurement setup.

Furthermore, as displayed in table 1, numerous participants were using psychotropic medication, most commonly antidepressants or stimulants. There are known effects of psychotropic medication on sleep architecture^40^, but after adjusting for psychotropic medication use, the results remained unchanged in terms of their direction and magnitude. Furthermore, none of the participants reported changing their psychotropic medication between the baseline measurement and three month-follow up. We therefore believe that the impact of psychotropic medication on our findings was small to minimal.

Altogether, our results show that sex hormones seem to change sleep architecture, but that the effects of sex hormones are dependent on one’s sex assigned at birth. Although these findings are from a relatively small population, they do provide evidence indicating there is a sex assigned at birth-specific predisposition for sensitivity to the effects of sex hormone fluctuations on sleep. This finding is important for research in transgender people, but it is also of major importance for studies on puberty, pregnancy and menopause. Sleep is an important component of well-being, and decreased sleep quality is associated with poorer physical and mental health^41^. If sex hormone exposure, during development or during adulthood, could predispose one to be more sensitive to poor sleep during sex hormone transitions, this is of interest for public health.

Future research should focus on studying underlying mechanisms in sleep and sex hormones, mainly on the effects of estradiol and testosterone in sleep-promoting regions of the human brain, as well as on longer-term effects of GAHT on sleep architecture. Furthermore, future research should assess whether the worsening in sleep architecture in transmasculine persons has effects on their well-being, most importantly on risk of depressive disorders.

## Data Availability

All data are available upon reasonable request.

## Data Availability

All data are available upon reasonable request.

## Acknowledgements

First and foremost, we would like to thank all the RESTED study participants for their participation and efforts, without which this study would not have been possible. Furthermore, we would like to thank all the interns and students who assisted in the data collection of the RESTED study: Rens Voskuijlen, Lotte van der Kolk, Claartje Binnerts, Jip Andriesse, Danique Hofs, Laura Varkevisser, Maureen González, Annefleur Zwager, Sigrid Theunis, Berend Slootman, Nina Pot and Isabel Löwensteijn. Lastly, we would like to thank the clinical staff of the Amsterdam UMC and University Medical Centre Groningen for their collaboration and efforts to help the study succeed.

