## Supplementary tables S1, S2 and S3. for "Influence of sex hormone use on sleep architecture in a transgender cohort: findings from the prospective RESTED study"

### Supplementary materials

#### 1. Hormone form and dosage

The table below displays the form and dosage of GAHT used by the study participants, at the start of GAHT (e.g. after the baseline measurement) and at the 3-month follow up.

**Table S1. Hormone formulation prescribed at the start of GAHT and 3 month follow-up**

| | | **Transmasculine participants (n = 38 )** | | **Transfeminine participants (n = 35)** | |
|---|---|---|---|---|---|
| **Measurement** | | Start of GAHT n = 36 | At 3 month follow-up n = 26 | Start of GAHT n = 32 | At 3 month follow-up n = 24 |
| **Cycle regulation use (n, %)** | Progestin only – oral | 4 (11%) | 3 (12%) | - | - |
| | Progestin only – non-oral methods [1.] | 8 (22%) | 3 (12%) | - | - |
| | Estradiol and progestin – combined oral contraceptives | 6 (17%) | 0 (0%) | - | - |
| | None | 18 (50%) | 6 (23%) | - | - |
| **Testosterone form (n, %)** | Transdermal | 32 (89%) | 19 (73%) | - | - |
| | Intramuscular – short-acting (esters) | 4 (11%) | 2 (8%) | - | - |
| | Intramuscular – long-acting (undecanoate) | 0 (0%) | 5 (20%) | - | - |
| **Estrogen form (n, %)** | Oral | - | - | 17 (53%) | 14 (46%) |
| | Transdermal – gel | - | - | 3 (9%) | 3 (13%) |
| | Transdermal – spray | - | - | 0 (0%) | 0 (0%) |
| | Transdermal – patches | - | - | 12 (38%) | 7 (29%) |
| **Anti-androgen form (n, %)** | GnRH analogues – short-acting | - | - | 25 (78%) | 13 (54%) |
| | GnRH analogues – long-acting | - | - | 4 (13%) | 8 (33%) |
| | Cyproterone actate | - | - | 3 (9%) | 3 (13%) |

[1.] Methods include hormonal intrauterine devices, intramuscular injections, progestin implants

#### 2. Smartsleep mode

The table below displays the outcomes stratified by sleep device setting and the estimated differences between the inactive and the active mode. Analyses show no significant differences between the active and inactive mode of the device.

| Table S2: sleep architecture during GAHT – with main predictor for devices in active mode. | | | | | | |
|---|---|---|---|---|---|---|
| | TM | | | TF | | |
| | Means and SD or median and IQR | | Estimated difference active vs. inactive mode [a.] | Means and SD or median and IQR | | Estimated difference active vs. inactive mode [a.] |
| **Setting** | *Inactive* | *Active* | | *Inactive* | *Active* | |
| **Measurement weeks** | Baseline: n = 33<br>3MO: n = 15 | Baseline: n = 3<br>3MO: n = 11 | Beta, 95% confidence interval, p-value | Baseline: n = 31<br>3MO: n = 23 | Baseline: n = 1<br>3MO: n = 1 | Beta, 95% confidence interval, p-value |
| **SOL (minutes)** | 17.8 (7.96 to 33.1) | 20.02 (7.96 to 36.7) | -15.5% (-39.1% to 17.7%)<br>P = 0.31 | 13.98 (7.29 to 27.25) | 16.87 (12.22 to 26.85) | 3.21% (-10.95 to 19.58)<br>P = 0.68 |
| **TST (hours)** | 7.3 (1.4) | 7.6 (1.6) | 0.144 (-0.30 to 0.59)<br>P = 0.53 | 6.49 (1.33) | 6.64 (0.67) | 0.6594 (-0.251 to 1.559)<br>P = 0.15 |
| **WASO (minutes)** | 20.27 (12.8 to 38.4) | 21.0 (15.4 to 28.1) | 4.2% ( -18.7 to 33.4) p = 0.75 | 15.79 ( 9.83 to 37.54) | 21.66 (11.18 to 32.1) | -9.998% ( -47.56 to 54.47)<br>P = 0.70 |
| **NRI** | 1 (0 to 2) | 1 (0 to 2) | 6.96% ( -10.4% to 27.8%)<br>P = 0.46 | 1 (0 to 1.5) | 1 (0 to 2) | -17.26% (-44.38 to 23.26)<br>P = 0.35 |
| **NRA** | 27 (21 to 40) | 26.5 (19 to 43) | -3.1% ( -17.6 to 13.9)<br>P = 0.70 | 27 (41 to 18) | 40 (15.5 to 53.5) | - 13.80% (-41.08 to 26.17)<br>P = 0.45 |
| **SWS (minutes)** | 87 (31) | 93 (32) | -1.5 (-5.5 to 8.59)<br>P = 0.67 | 86 (30) | 82 (23) | -6.31 (-21.53 to 8.96)<br>P = 0.42 |
| **% SWS** | 20.0% (7.3%) | 21.0% (8.0%) | -0.04% ( -1.88 to 1.80)<br>P = 0.95 | 22.6 (8.05) | 20.82 (7.02) | -4.3247% (-9.2956 to 0.692)<br>P = 0.091 |
| **REM sleep duration (minutes)** | 118.9 (45.5) | 119.1 (38.9) | -9.90 (-23.27 to 3.49)<br>P = 0.149 | 114.26 (42.9) | 106.91 (19.67) | 7.938 (-20.63 to 36.28)<br>P = 0.58 |

| | | | | | | |
|---|---|---|---|---|---|---|
| **REM sleep latency (minutes)** | 82 (33.5 to 125.5) | 68 (28.8 to 101.5) | -1.9% (-29.6 to 39.3)<br>P = 0.95 | 81.00 (22.5 to 111.5) | 89.0 (39.5 to 95.0) | 29.74% (- -42.66 to 90.51)<br>P = 0.53 |
| [a.] Analyzed using a linear mixed model with measurement phase and device setting as fixed predictors and a random intercept per participant, with inactive mode as the reference category. | | | | | | |

#### 3. Participation and non-participation in objective sleep measurements

The table below displays demographic descriptions and questionnaire outcomes of participants who contributed sleep device measurements to the final dataset and participants who did not. It is possible that participants took part in the objective sleep measurements, but that the measurements were of low quality and that they were excluded, which means the participant's measurements are therefore not present in the final dataset.

**Table S3. Demographic and clinical characteristics of participants opting in and out of sleep device measurements. IQR = Interquartile range.**

| | | Not present in objective sleep measurements | Present in baseline objective sleep measurements only | Present in 3MO objective sleep measurements only | Present in baseline and 3MO |
|---|---|---|---|---|---|
| **n** | | 26 | 23 | 5 | 45 |
| **Age** | *Median, IQR* | 22 (20 to 24.5) | 24 (21.5 to 26) | 28 (19 to 30) | 24 (22 to 27) |
| **Group (n, %)** | *TM* | 13 (50%) | 12 (52%) | 2 (40%) | 24 (53%) |
| | *TF* | 13 (50%) | 11 (48%) | 3 (50%) | 21 (47%) |
| **Psychotropic medication use (n, %)** | *Yes* | 5 (19%) | 2 (9%) | 1 (20%) | 9 (20%) |
| | *No* | 21 (81%) | 21 (91%) | 4 (80%) | 36 (80%) |
| ***Scores at baseline*** | | | | | |
| **ISI scores (range 0 to 28)** | *Median*<br>*IQR*<br>*missings (n)* | 7.5<br>4.75 to 10.25<br>2 | 5.5<br>3.25 to 10.5<br>1 | 6.0<br>4.0 to 6.0<br>0 | 6.0<br>3.0 to 9.0<br>0 |
| **PSQI scores (range 21)** | *Median*<br>*IQR*<br>*missings (n)* | 7<br>4.25 to 9.0<br>4 | 5<br>4 to 9<br>4 | 4<br>4 to 6<br>0 | 6<br>4 to 8<br>4 |
| **IDS-SR scores (range 0 to 84)** | *Median*<br>*IQR*<br>*missings (n)* | 15.5<br>10 to 26<br>4 | 12<br>8 to 25.75<br>1 | 9<br>8 to 25<br>0 | 12<br>8 to 21<br>0 |
| **PSS scores (range 0 to 40)** | *Median*<br>*IQR*<br>*missings (n)* | 13<br>9.5 to 16.25<br>2 | 12<br>8.25 to 17<br>1 | 11<br>6 to 13<br>0 | 10<br>8 to 16<br>0 |
